# SARS-CoV-2 antibody prevalence in Sierra Leone, March 2021: a cross-sectional, nationally representative, age-stratified serosurvey

**DOI:** 10.1101/2021.06.27.21259271

**Authors:** Mohamed Bailor Barrie, Sulaiman Lakoh, J. Daniel Kelly, Joseph Sam Kanu, James Squire, Zikan Koroma, Silleh Bah, Osman Sankoh, Abdulai Brima, Rashid Ansumana, Sarah A. Goldberg, Smit Chitre, Chidinma Osuagwu, Justin Maeda, Bernard Barekye, Tamuno-Wari Numbere, Mohammed Abdulaziz, Anthony Mounts, Curtis Blanton, Tushar Singh, Mohamed Samai, Mohamed A. Vandi, Eugene T. Richardson

**Affiliations:** Institute for Global Health Sciences, University of California, San Francisco, USA; Partners In Health, Sierra Leone; Department of Epidemiology and Biostatistics, University of California, San Francisco, USA; Ministry of Health and Sanitation, Sierra Leone; Statistics Sierra Leone; Njala University, Sierra Leone; School of Public Health, Faculty of Health Sciences, University of the Witwatersrand, Johannesburg, South Africa; Heidelberg Institute of Global Heath, University of Heidelberg Medical School, Germany; Department of Global Health & Social Medicine, Harvard Medical School, Boston, MA, USA; Lewis Katz School of Medicine at Temple University, Philadelphia, PA, USA; Africa Centres for Disease Control and Prevention, Addis Ababa, Ethiopia; U.S. Centers for Disease Control and Prevention, Atlanta, GA, USA; College of Medicine and Allied Health Sciences, Freetown, Sierra Leone; Department of Medicine, Brigham and Women’s Hospital, Boston, MA, USA

## Abstract

**Background:** As of 26 March 2021, the Africa CDC had reported 4,159,055 cases of COVID-19 and 111,357 deaths among the 55 African Union Member States; however, no country has published a nationally representative serosurvey as of May 2021. Such data are vital for understanding the pandemic’s progression on the continent, evaluating containment measures, and policy planning.

**Methods:** We conducted a cross-sectional, nationally representative, age-stratified serosurvey in Sierra Leone in March 2021 by randomly selecting 120 Enumeration Areas throughout the country and 10 randomly selected households in each of these. One to two persons per selected household were interviewed to collect information on socio-demographics, symptoms suggestive of COVID-19, exposure history to laboratory-confirmed COVID-19 cases, and history of COVID-19 illness. Capillary blood was collected by fingerstick, and blood samples were tested using the Hangzhou Biotest Biotech RightSign COVID-19 IgG/IgM Rapid Test Cassette. Total seroprevalence was was estimated after applying sampling weights.

**Findings:** The overall weighted seroprevalence was 2.6% (95% CI 1.9-3.4). This is 43 times higher than the reported number of cases. Rural seropositivity was 1.8% (95% CI 1.0-2.5), and urban seropositivity was 4.2% (95% CI 2.6-5.7).

**Interpretation:** Although overall seroprevalence was low compared to countries in Europe and the Americas (suggesting relatively successful containment in Sierra Leone), our findings indicate enormous underreporting of active cases. This has ramifications for the country’s third wave (which started in June 2021), where the average number of daily reported cases was 87 by the end of the month—this could potentially be on the order of 3,700 actual infections, calling for stronger containment measures in a country with only 0.2% of people fully vaccinated. It may also reflect significant underreporting of incidence and mortality across the continent.

**Funding:** This study was supported by NIAID K08 AI139361, the Sierra Leone Ministry of Health and Sanitation, and the Africa CDC.

## Background

As of 26 March 2021, the Africa CDC had reported 4,159,055 cases of COVID-19 and 111,357 deaths among the 55 African Union Member States;^1^ no African Union (AU) Member States has published a nationally representative COVID-19 antibody serosurvey as of May 2021.

Sierra Leone reported 3,962 cases (49.5 per 100,000 population) and 79 deaths as of 26 March 2021 (Figure 1), with 2.9% of all RT-PCR tests becoming positive. This relatively low case rate compared with countries in Europe and the Americas could be partly explained by the fast implementation of stay-at-home measures, collaboration at regional levels, and a swift response at the continental level.^2^ Indeed, Ministers of Health and the Africa CDC convened to develop a continental strategy early in the pandemic.^3,4^

**Figure 1.**
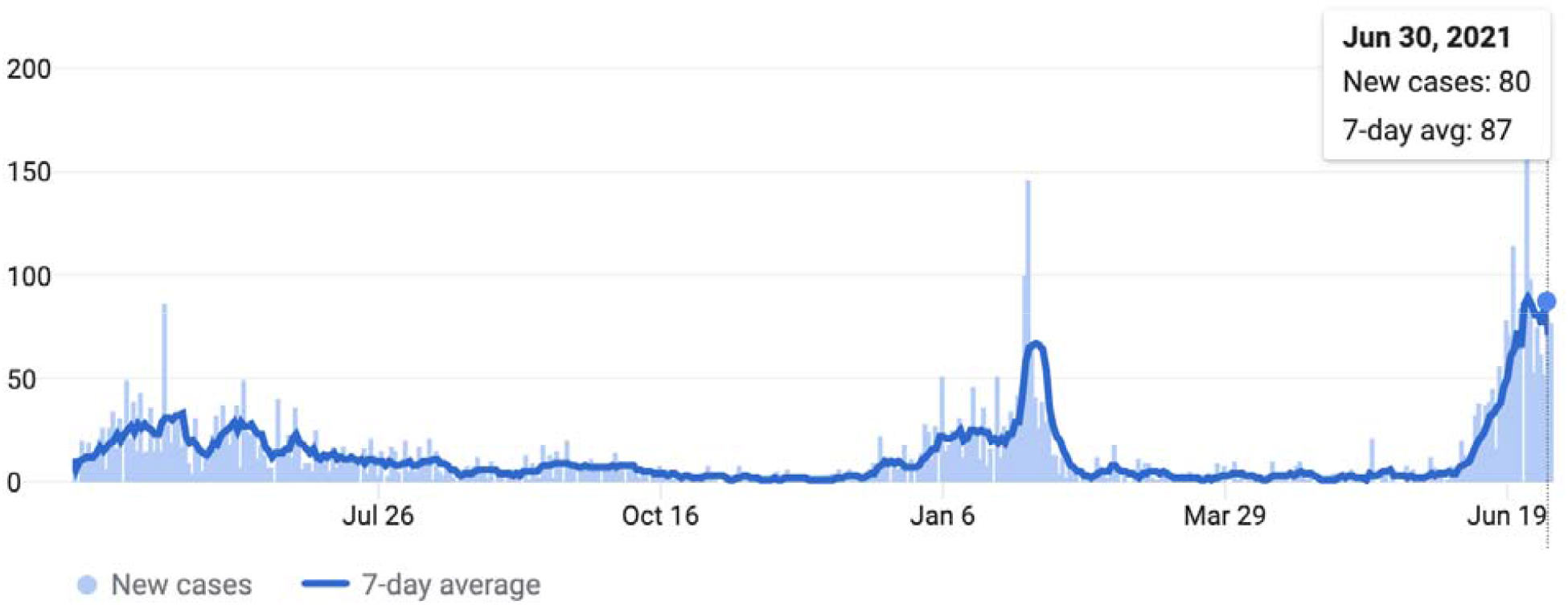
Daily COVID cases in Sierra Leone until from Feb 1, 2020 to June 30, 2021. The country’s first case was reported on March 31, 2020. (Source: COVID-19 Dashboard by the by the Center for Systems Science and Engineering at Johns Hopkins University. Accessed July 2, 2021).

To date, however, all reported data on cases and deaths have come from event-based surveillance.^5^ Case reporting is influenced by often under-resourced strategies for case finding, testing, and contact tracing, and might underestimate the true burden of severe acute respiratory syndrome coronavirus 2 (SARS-CoV-2) infection.^6^ We therefore sought to measure country-wide seroprevalence of SARS-CoV-2 antibodies in Sierra Leone, an AU Member State, as such data are vital for understanding the pandemic’s progression in the country and on the continent, as well as for evaluating containment measures and policy planning.^7^

## Methods

### Design

This was a cross-sectional, population-based, age-stratified serosurvey, targeting household members aged 5 years or above, regardless of previous or current infection with COVID-19, who resided in Sierra Leone during the period of transmission of SARS-CoV-2 (that is, since the first case was reported on March 31, 2020).

For the purpose of sample size estimation, seroprevalence was estimated at 5%. Considering 95% confidence intervals and a design effect of 3, the minimum sample size was calculated to be 1,200 households with at least one member of each household selected. This was designed to give a total of 240 individuals for each of the age strata 5-9, 10-19, 20-39, 40-59 and ≥60 years old.

### Sampling

The sampling frame was the most recent census conducted by Statistics Sierra Leone.^8^ We conducted randomized, multistage sampling, with the first stage consisting of 120 randomly chosen Enumeration Areas (EAs), which are small units that contain 80-120 households each. Within each selected EA, households were identified on a satellite map by numbering them in order west to east, then north to south. After that, 10 households were chosen using a random number generator. One to two members of each household over the age of 5 years were then selected for participation, and the selection of individuals was constrained to give a comparable number of individuals for each age stratum: In Sierra Leone, approximately 41% of the population is aged ≤ 15 years and only about 3.5% are aged ≥ 65 years.^8^ This means that any straight-forward random sample of the population will result in an insufficient sample of older adults. For example, a truly random sample of the population that selected 1,000 people, would include 410 children aged ≤ 15 years and only 35 adults aged ≥ 65 years. This would result in a sample too small to give confidence in the estimated seroprevalence of senior adults. To compensate for this, the sampling process was constrained to restrict the number of children selected, and adults were over-sampled.

We started by selecting individuals from the older strata in a systematic progression from house-to-house. In this method, the survey team would attempt to capture an individual of each sex in the older strata before progressing to younger strata. If there was not any individual available for the appointed age group, they would skip to the next younger age stratum and choose from that one.

### Test Validation and Sample collection

Fingerstick samples were screened for the presence of both IgM and IgG SARS-CoV-2 virus specific antibodies using the Hangzhou Biotest Biotech RightSign COVID-19 Rapid Test Cassette, which in FDA testing had combined IgM/IgG specificities of 100% and IgM/IgG sensitivities of 100%.^9^ We further validated the test with a control panel of 58 serum samples (from 18 persons tested by NP swab—10 who tested negative and 8 positive, with 5 serial dilutions of each positive) and also found combined IgM/IgG sensitivities and specificities of 100%.

All tests were performed at the time of interview and according to manufacturer’s instructions. Each participant was notified of their results during the interview. Participants with positive IgM results were referred to the District Health Management Team as an active case.

### Data analysis

We collected demographic data on study participants including age, sex, district, number of people per household, occupation, and whether they lived in a rural or urban area (those residing in district capitals were categorized as urban, and the rest as rural). As in other surveys, we categorized the reported occupations into high-risk and low-risk categories on the basis of the risk of exposure to potential COVID-19 cases.^6^ For example, occupations such as health-care workers, shopkeepers or petty traders, transport operators, and those in food service were considered high-risk occupations; on the other hand, farmers, miners, homemakers, students, were considered as being at lower risk of exposure. The information about occupation of the participants was captured as open-ended text and was categorized into high and low risk by the investigators.

The data were analyzed to estimate an unadjusted seroprevalence of IgM/IgG antibodies against SARS-CoV-2 with 95% CI. An individual was deemed seropositive if they were IgM+/IgG-, IgM-/IgG+, or IgM+/IgG+. We also calculated seroprevalence by age group, sex, and area of residence (rural/urban). To estimate the weighted seroprevalence, we post-stratified by district (counting Falaba as part of Koinadugu) and calculated sampling weights as a ratio of the national population to district population (aged >5 years). We then dtermeined if there were significant differences in seropostivity by demographic variable using the Rao-Scott Chi-square test.

### Ethics Approval

The study was approved by the Sierra Leone Ethics and Scientific Review Committee. It received a Not Research Determination (IRB20-1394) from the Harvard Institutional Review Board as it was deemed Public Health Surveillance.

## Findings

In March, 2021, we identified 1,893 individuals aged 5 years or older from households in 120 EAs. Three hundred forty (18%) of the participants were age 5-9; 384 (20.3%) 10-19; 451 (23.8%) 20-39; 359 (19.0%) 40-59; and 359 (19.0%) >60. Slightly more than half of the participants (964, 50.9%) were female; 1,174 (62%) were residing in rural areas, and 845 (44.6%) had an occupation with a relatively higher risk of exposure to people potentially infected with COVID-19. No individuals reported having previously tested positive for COVID-19 (Table 1).

**Table 1:** Participant characteristics.

|  | Participants<br>(n=1,893) |
| --- | --- |
| Age, years |  |
| < 10 | 340 (18.0%) |
| 10-19 | 384 (20.3%) |
| 20-39 | 451 (23.8%) |
| 40-59 | 359 (19.0%) |
| > 60 | 359 (19.0%) |
| Gender |  |
| Female | 964 (50.9%) |
| Male | 929 (49.1%) |
| Area of residence |  |
| Rural | 1,174 (62.0%) |
| Urban | 719 (38.0%) |
| Number of People Per Household<br>(n = 1,719)<sup>§</sup> | 7 (5-9)*<br>(1-45)<sup>†</sup> |
| Occupation with high risk of exposure to<br>COVID-19<br>(n = 1,892)<sup>§</sup> | 845 (44.6%) |
| Current smoker<br>(n = 1,892)<sup>§</sup> | 250 (13.2%) |
| Would accept vaccine if offered<br>(n = 1,892)<sup>§</sup> | 1,666 (88.0%) |
| Seropositive | 53 (2.8%) |
| Positive IgM | 12 (0.6%) |
| Positive IgG | 44 (2.3%) |
| * median (interquartile range) |  |
| † (absolute range) |  |
| § Data are n (%) unless otherwise stated |  |
| **Table 1:** Participant characteristics. |  |

The overall weighted and adjusted seroprevalence was 2.6% (95% CI 1.9-3.4). IgM positivity was 0.6% and IgG positivity was 2.3%; only 6 participants (0.3%) were IgM+/IgG+. Rural weighted seropositivity was 1.8% (95% CI 1.0-2.5) and urban seropositivity was 4.2% (95% CI 2.6-5.7). Stratifying by age group and weighting, 1.7% (95% CI 0.2-3.2) of participants age 5-9 tested positive for anti-SARS-CoV-s antibodies, as did 2.6% (95% CI 0.8-4.2) of those 10-19, 1.2% (95% 0.2-2.3) of those 20-39, 4.4% (95% CI 2.4-6.4) of those 40-59, and 3.6% (95% CI 1.6-5.6) of those 60 and above (Table 2). None of the 35 healthcare workers sampled tested positive. There was a significant difference in seropositivity between rural and urban populations (Rao-Scott Chi-square p=0.002). Seropositivity in females was 74% higher than in males, nearly reaching significance (Rao-Scott Chi-square p=0.056).

**Table 2:**
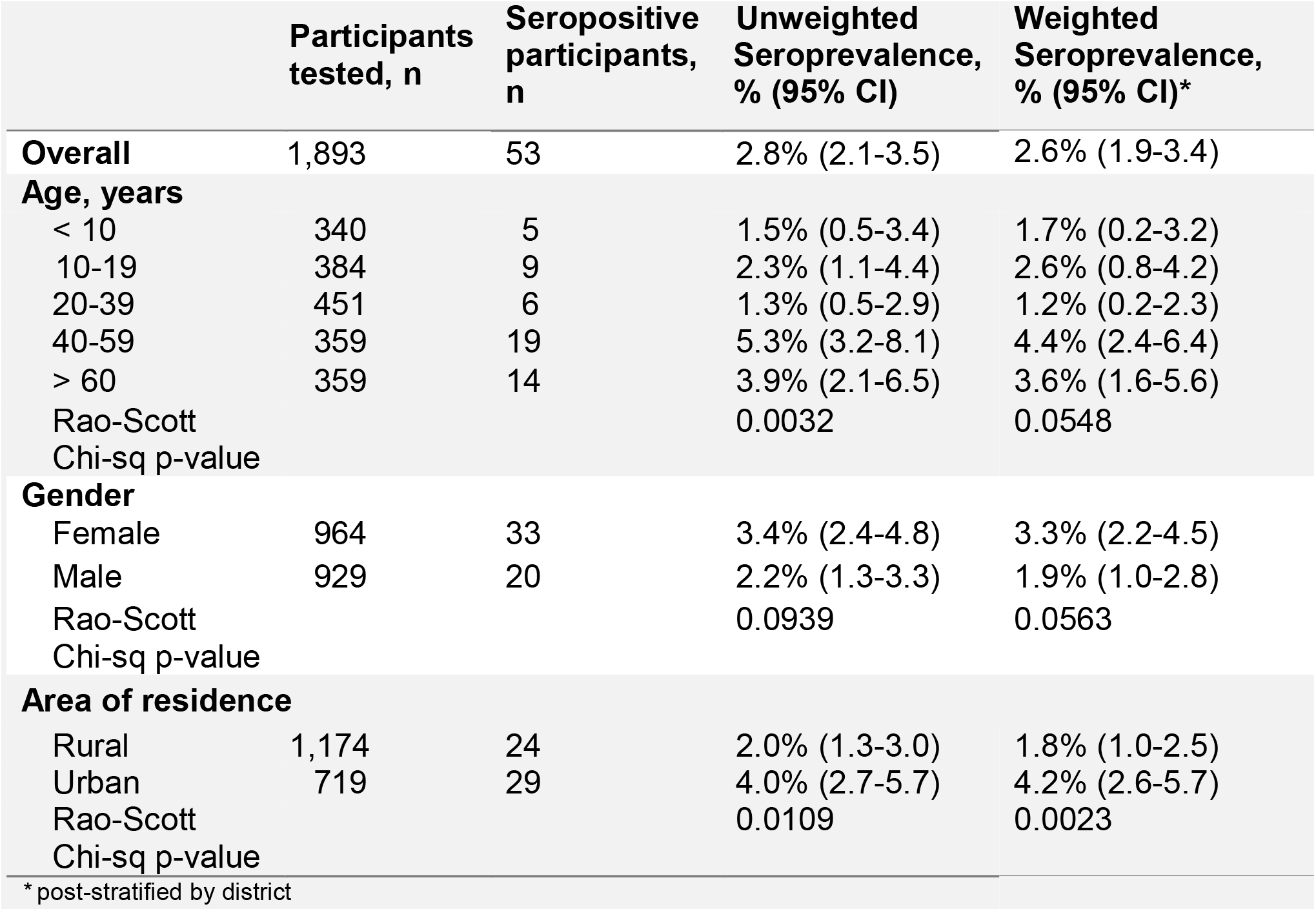
Seroprevalence by demographic characteristics.

## Interpretation

Our findings indicate that 2.6% of Sierra Leone’s population aged 5 years or older had been infected with SARS-CoV-2 infection by March 2021, with an estimated 203,060 infections. This is 43 times higher than the reported number of cases, which may give an idea of the potential disease burden as of June 30, 2021, where the average number of daily reported cases was 87 (represeting the onsent of the third wave in the country, Figure 1).^3^ Furthermore, this finding demonstrates that, similar to many other countries,^10^ herd immunity is far from being reached in Sierra Loene. Notably, seroprevalance in urban settings was more than double that of rural. This trend has been seen in other surveys and can be explained by higher contact rates and challenges in safe physical distancing in urban areas.^6^

While a total seroprevalence of 2.6% is significantly lower than city and regional estimates across the continent, it represents the first nationally representative SARS-CoV-2 serosurvey to be published by an African nation. (Of note, West Africa was one of the last regions of the world to be affected by H1N1 influenza in 2009-2010,^11^ so a similar dynamic may be occurring here.)

Chibwana and colleagues found that 12.3% of healthcare workers in Blantyre, Malawi tested positive for SARS-CoV-2 antibodies;^12^ Uyoga and colleagues found a crude seroprevalence of 5.6% in Kenyan blood donors;^13^ Olayanju and colleagues determined a 45.1% seroprevalence among asymptomatic healthcare workers in Ibadan, Nigeria;^14^ and Mulenga and colleagues noted a SARS-CoV-2 prevalence of 10.6% in 6 of 117 districts in Zambia.^15^

Significant underreporting of cases may be the result of a high prevalence of minimally symptomatic disease in a younger demographic coupled with under-resourced systematic surveillance in the setting of legacies of distrust of authorities and foreign interventions (resulting in the avoidance of testing).^16–24^ Indeed, Mwananyanda and colleagues found significant underreporting of cases in Zambia through postmortem surveillance.^25^ Furthermore, 1) limited testing capacity, 2) low acquired immunity to SARS-CoV-2, 3) and less than 0.2% of Sierra Leoneans being fully vaccinated as of 21 May 2021 is extremely concerning in that it presents a very large population of susceptible individuals at risk for future variant waves. As reported by Reuters, during the first week of May 2021, Sierra Leone averaged about 556 doses of vaccine administered each day: At that rate, it would take 2,809 days (7.7 years) to administer enough doses for 10% of the population.^26^ Indeed, both under-reporting of cases and limited vaccination campaigns increase the risk of new variants emerging and circulating in the population before public health authorities have a chance to detect them and prevent their spread.^27^ Since India was forced to halt the export of vaccines on account of the devastating second wave there, this timeline could worsen.^28^ These data should therefore strengthen demands for faster vaccine deployments in countries like Sierra Leone as part of global vaccine justice, including support for intellectual property waivers and technology transfers.^29^

Our study has two notable limitations. The effects of waning immunity on the performance of the assay we used is unclear, and we therefore may not have fully captured those individuals infected in the first wave of COVID-19 in spring 2020 (Figure 1).^30^ In addition, the exact sensitivity of the assay for detecting minimally symptomatic infection is not known. Both factors would bias the estimated seroprevalence downwards.

## Data Availability

A subset of the key anonymized individual participant data collected is available upon request to the corresponding author, after approval of a proposal with a signed data
access agreement.

## Acknowledgements

We would like to thank Yelena Gorina for her help with data analysis.

